# Serological Survey in a university community after the fourth wave of COVID-19 in Senegal

**DOI:** 10.1101/2024.01.28.24301904

**Authors:** Fatou Thiam, Abou Abdallah Malick Diouara, Clemence Stephanie Chloe Anoumba Ndiaye, Ibrahima Diouf, Khadim Kebe, Assane Senghor, Djibaba Djoumoi, Mame Ndew Mbaye, Idy Diop, Sarbanding Sane, Seynabou Coundoul, Sophie deli Tene, Mamadou Diop, Abdou Lahat Dieng, Mamadou Ndiaye, Saidou Moustapha Sall, Massamba Diouf, Cheikh Momar Nguer

## Abstract

Cross-sectional survey was conducted to assess the true extent of COVID-19 exposure among students and staff of Polytechnic High School (PHS). Random cluster sampling was carried out between May 19 and August 18, 2022, after the fourth wave of COVID-19 transmission. IgM and IgG SARS-CoV-2 antibodies were screened using WANTAI SARS-CoV-2 ELISA assays. Seroprevalence and descriptive statistics were calculated. Moreover, the associations between seropositivity and different factors (age, gender, preventive, etc.) were determined using logistic regression. A total of 637 participants were recruited, including 563 students, followed by 52 TAS officers, and 22 professors. The median age among was 21 years [18–63]. 62.0% of the participants were female, and 36.89% were male, with a male-to-female ratio = 0.59. The overall IgG and IgM seroprevalence were 92% and 6.91% respectively. We found a higher IgM seroprevalence in men than women (9.4% vs. 5.6%) and a lower IgM seroprevalence in (18-25) age group compared to (55-65) years. Low compliance with preventive measures was found with a significant IgM seroprevalence depending on non-respect of social distancing (p = 0.008). IgM seropositivity was associated with Body Mass Indice (BMI) categorized (O.R. 0.238, *p* = 0.043), ethnic group (O.R. 0.723, *p* = 0.046) and marital status (O.R. 2.399, *p* = 0.021). Moreover, IgG seropositivity was associated with vaccination status (O.R. 4.741, *p* < 0.001). Our study revealed that the majority of students and staff have already been exposed to SARS-CoV-2, which confirms the circulation of SARS-CoV-2 in PHS at the time of the survey. Our results underline the importance of seroepidemiological surveys to estimate the real impact of the COVID-19 pandemic in a community and to monitor disparities in antibody response in the population.

## Introduction

The Coronavirus 2019 (COVID-19), newly identified in December 2019 in Wuhan, China, is an infectious disease caused by Severe Acute Respiratory Syndrome Coronavirus 2 (SARS-CoV-2) [1, 2]. On January 30 2020, the World Health Organization (WHO) declared COVID-19 a public health emergency of global concern, and on March 11, 2020, it announced the COVID-19 epidemic as a pandemic [3]. By November 12, 2023, more than 697 million of people had been infected worldwide, with over 6.9 million deaths [4].

The COVID-19 outbreak had substantial economic, social, and health repercussions and damaging educational consequences [5–7]. In April 2020, the World Bank estimated that higher education institutions had been closed in 175 countries and that studies had been interrupted or significantly disrupted due to COVID-19, affecting more than 220 million students [8]. The pandemic forced academic communities to adopt online platforms for the continuity of teaching and learning activities, negatively impacting learning outcomes, particularly in developing countries where the lack of network infrastructures, computers, and internet access is challenging distance learning in developing countries [9, 10].

In Senegal, a West African country, the first case of SARS-CoV-2 was identified on March 2, 2020. Since then, the number of cases has risen considerably, and the country currently has more than 89,022 confirmed cases of COVID-19 and 1,971 deaths [11]. The onset of the pandemic led the government authorities to close higher education establishments from March 16 to August 31 2020. The reopening of the establishments took place with the introduction of sanitary protocols that emphasized the reinforcement of barrier measures (wearing masks, hand washing with soapy water or hydroalcoholic gel, respect for social distancing and systematic taking of body temperature at campus access points) [12]. However, these measures have often been insufficient to stem the spread of the virus. Clusters quickly appeared in some establishments, including the Cheikh Anta Diop (CAD) University, located in Dakar, the capital of Senegal.

The failure of strategies to combat the spread of the virus could be linked to multiple factors, including (i) CAD University ecosystem, with its over 80,000 students from diverse origins. The consequence is overcrowding in student residences that caused the spread of the virus; (ii) the university environment is mainly populated by young people, who do not seem to be fully aware of the risks associated with the COVID-19 health crisis [13]; (iii) the promiscuity in the Halls of residence, making it difficult to comply with social preventive measures; (iv) Lastly, lack of vaccine deployment and reluctance was noted among young people, which also contributes to low vaccination coverage in universities [14].

Faced with all these difficulties, it was essential to adopt cyclical measures to manage the COVID-19 pandemic, characterised by the virus spreading in waves [15, 16]. These measures could involve introducing survey studies based on diagnostic testing and mass screening protocols to diagnose and follow up with people exposed to or infected with SARS-CoV-2. In practice, detecting viral RNA by RT-PCR is the reference method for confirming the diagnosis of SARS-CoV-2 infection [17, 18]. However, access to diagnostic tests still needs to be improved in poor countries [19]. The other way to estimate the true extent of the epidemic is to conduct Seroprevalence surveys [20, 21]. Seroepidemiological studies to detect the presence of anti-SARS-CoV-2 antibodies are a valuable tool for assessing the timing of the epidemic. It can help to confirm the presence of a recent infection when PCR is limited. Some antibodies, such as IgG, can even be detected years after exposure [22]. Their detection can be used to identify previous or recent exposure to SARS-CoV-2. By carrying out this analysis on a representative population, it is possible to estimate what proportion of the population has already been exposed to the new coronavirus [23]. This information is helpful in identifying the epidemic phases and can help the authorities make decisions and even anticipate appropriate measures to contain the pandemic’s spread [24]. Sero-epidemiological studies have been conducted in academic institutions such as universities in many countries [25–28]. However, more studies are highly relevant because university communities (faculty, staff and students) could be among the most exposed to SARS-CoV-2. In Senegal, no seroprevalence studies have been conducted in cohorts of educational institutions.

Therefore, we conducted an on-site screening project at Polytechnic High School (PHS), a public institution with an inter-African vocation at CAD University from May to August 2022 after the fourth wave of Coronavirus disease in Senegal. We aimed to assess the true extent of previous and recent COVID-19 exposition among students and staff and to investigate the risk factors associated with SARS-CoV-2 IgM and IgG seropositivity.

## 2. Materials and Methods

### 2.1. Study Design and Population

The SARSESP (“Etude de Seroprevalence du SARS-CoV-2 au sein de l’École Supérieure Polytechnique”, in the Cheikh anta DIOP University (CAD University)) project is an on-site university population-based cross-sectional study. The sampling occurred at the PHS, an establishment of CAD University. Students, Professors, and Technicians, Administrative and Service (TAS) officers at SPS were invited by e-mail to enrol in the study. Participants were volunteers who registered online between 19 May and 18 August 2022. A questionnaire was administered to each participant after consent, blood samples were taken for SARS-CoV-2 antibody detection.

### 2.2. Sample Size Calculation

We used stratified random sampling: the first stratum concerned professors, the second TAS officers and the third students. Systematic random sampling was used within each stratum to determine the required number of subjects. Using the lists provided by the student affairs and human resources departments, we calculated the sample size based on the hypothesis of an expected seroprevalence of 45%, with a precision of 5%, a design effect of 1.96, and a nonresponse rate of 65%. We determined that >450 participants needed to be recruited. To increase the accuracy of the results, this size was multiplied by 2, giving a sample size of 898, and rounded up to 1000 to account for any lost records. The survey step for selecting statistical units was 6133/1000, i.e. a step = 6. In this study, an allocation proportional to the size of each stratum was used. Thus, we considered a proportion of 83.54% for students, 9.97% for professors and 6.47% for TAS officers. Then, we obtained 65 TAS officers, 100 professors and 835 students. The inclusion criteria were age over eighteen (18) years, informed consent signed, and questionnaire completion. Then, the non-inclusion criteria were under 18 years of age, non-consent and contraindications to venous blood sampling (anaemia, Infection or hematoma at a prospective venipuncture site, etc.).

### 2.3. Questionnaire

We shared an interviewer-administered questionnaire with participants on an electronic tablet (https://enquete.ucad.sn/index.php/598685?lang=fr). The questionnaire covered points regarding different factors to assess for relationships between IgM and IgG seropositivity and these factors. Questions relating to sociodemographic characteristics were collected, such as age, sex, occupation, education level, nationality, ethnic group and accommodation type. We also collected alcohol and tobacco intake, SARS-CoV-2 vaccination status, and preventive measures related to SARS–CoV–2 practices. We provided all recruitment participants with face masks and hand sanitisers and encouraged them to practice physical and social distancing. The questionnaire on COVID-19 was posted online by the IT and Information Systems Department of CAD University.

### 2.4. Blood Collection and SARS-CoV-2 Antibodies Detection

After each participant signed the written consent form, a 10 mL whole blood sample was collected into a dry vacutainer tube by standard venipuncture technique. Blood samples were centrifuged at 2,500 rpm for 10 minutes. Then, the plasma was collected and stored in cryotubes at -80°C at the GRBA-BE laboratory biobank in PHS until the tests were carried out. Seropositivity to anti-SARS-CoV-2 antibodies was used as a biomarker of exposure to the SARS-CoV-2 virus. IgM class immunoglobulins were used as a marker of recent or current infection (usually from days 5-7 after symptoms appear, but sometimes later and decrease at days 15-22), and IgG class immunoglobulins were used as a biomarker of older infection (detectable from day 11 post symptom, reaching a maximum 3-4 weeks after and persist days 90 after) [29, 30]. Serological tests were performed by qualitative ELISA for IgM and quantitative ELISA for IgG following the instructions for the WANTAI SARS-CoV-2 IgM ELISA (Beijing Wantai Biological Pharmacy Enterprise, Beijing, China; Ref. WS-1196 and WS-1396) recommended by the WHO for seroepidemiological studies, which detects total antibodies (including IgM and IgG) binding the SARS-CoV-2 spike protein receptor binding domain (S1/RBD) [31]. Serum samples were analysed in duplicate according to the supplier’s recommendations.

### 2.5. Statistical Analyses

Statistical analyses were performed using Rstudio (version R 4.2.1) and GraphPad Prism (version 10.1.1 (316)) softwares. Continuous variables were described as mean (standard deviation) or median (interquartile range). Normally distributed variables were compared with a t-test, and nonparametric data were compared with the Mann-Whitney test. Categorical variables were presented as percent, and Fisher exact tests or chi-squared tests were used for proportional assessments. For all statistical tests, we accepted a two-sided level of significance was set at p ≤ 0.05.

SARS-CoV-2 seroprevalence was defined as the ratio of the number of people who developed anti-SARS-CoV-2 antibodies to the general population. Confidence intervals (95% CI) for seroprevalence were estimated using the Clopper-Pearson method. Using logistic regressions, we sought to understand how the IgM and IgG seropositivity are influenced by various variables such as age, gender, occupation, education level, nationality, type of accommodation, etc. The analyses were carried out after carefully handling the database and processing missing data.

### 2.6. Ethics Statement

This research complies with ethical recommendations. The project protocol has been validated by the National Health Research Ethics Committee (CNERS) of the Ministry of Health and Social Action. The research protocol was drawn up in accordance with Senegalese laws and regulations governing the confidentiality of personal data. The study was approved by the Senegalese National Ethics Committee for Research in Health (Reference number N°000043/MSAS/CNERS/SP, 28 February 2022).

## 3. Results

### 3.1. Baseline characteristics of participants

From the 1000 students, professors and TAS officers of PHS who were invited to participate in the study between 19 May 2022 and 18 August 2022, 637 (63.7%) participants were finally included (Figure 1). The most represented group was students (88.38%), followed by TAS officers (8.16%), and professors (3.45%). The mean age was 23.20 years, ranging from 18 to 63 years. The population’s median age was 21 years, with most participants aged 18 to 25 years, representing 85.7% of the population (Table I). 62.0% of the participants were female, and 36.89% were male, with a male-to-female ratio = 0.59. Most participants were Senegalese (94.66%) and lived in family homes (54.6%). In addition, most ethnic groups in the PHS population were Wolof (31.24%), Fula (24.33%) and Serer (21.98%). The prevalences of active smoking and taking alcohol were relatively low in the PHS community at the moment of the survey (1.9%) and (3.62%), respectively. Demographic data are shown in Table I. Calculation of body mass index also revealed the highest prevalence of normal weight (37.68%), followed by underweight (10.8%) and overweight/obesity, 8.32%). The most common blood group is O+ (47.08%). At the time of data collection, unvaccinated *vs.* vaccinated participants were 57.46% and 35.01 %) respectively.

**Figure 1.**
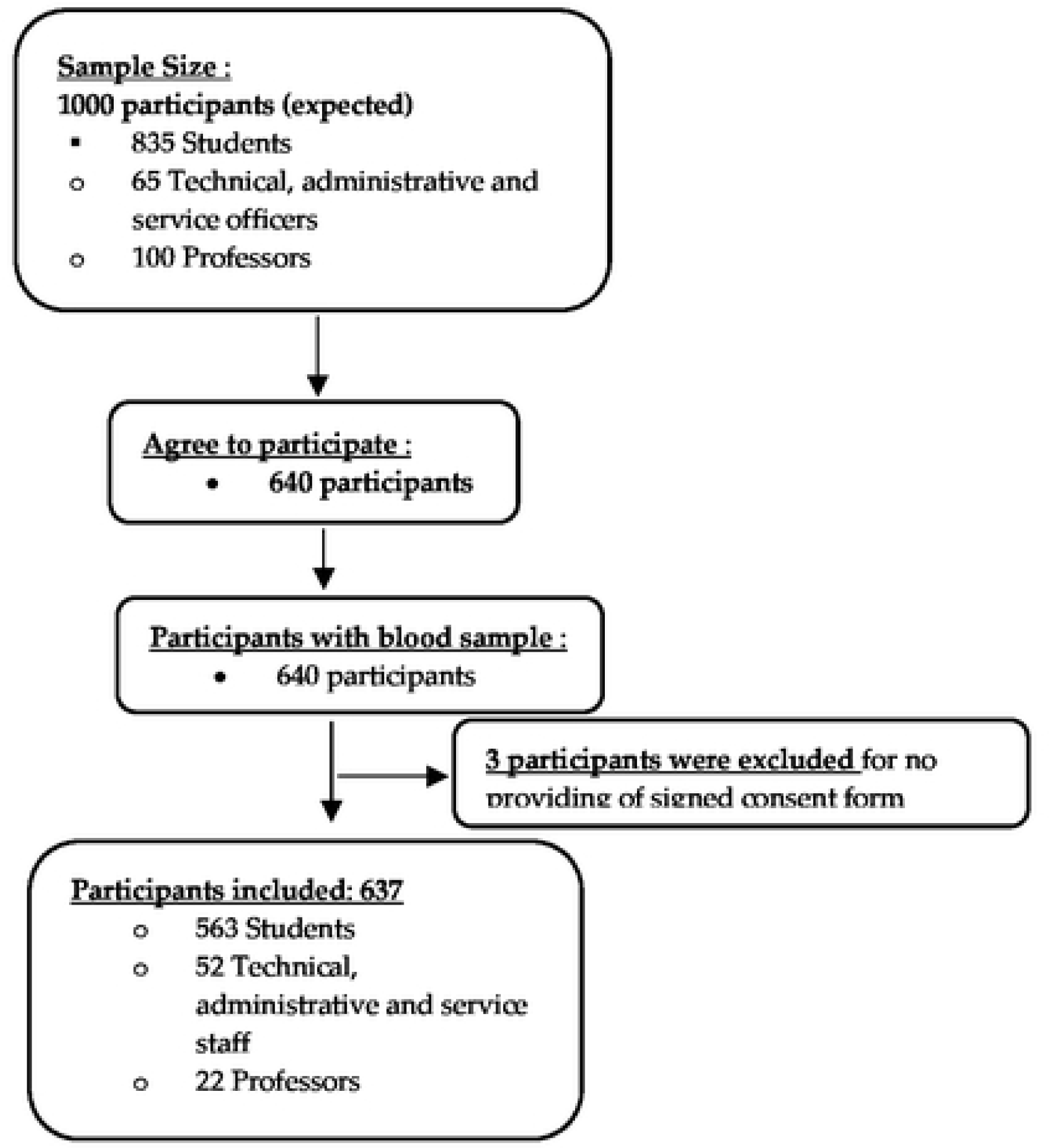
Flowchart of participants’ enrolment for the anti-SARS-CoV-2 antibody seroprevalencestudy in Superior Polytechnic School

**Table I:** Baseline characteristics of the study population.

| **Variable** | **Value, N (%)** |
| --- | --- |
| **Overall** | 637 (100) |
| **Age (years)** |  |
| Mean | 23.20 |
| Median | 21 |
| Range | 18 - 63 |
| **Age group (years)** |  |
| [18-25] | 546 (85.71) |
| (25-35] | 38 (5.96) |
| (35-45] | 26 (4.08) |
| (45-55] | 12 (1.88) |
| (55-65] | 6 (0.94) |
| **Gender** |  |
| Female | 395 (62) |
| Male | 235 (36.89) |
| **Occupation** |  |
| Students | 563 (88.38) |
| Professors | 22 (3.45) |
| TASO | 52 (8.16) |
| **Level of education** |  |
| Bachelor degree | 450 (70.64) |
| Master degree | 117 (18.36) |
| PhD | 20 (3.14) |
| others | 18 (2.82) |
| **Nationality** |  |
| Senegalese | 603 (94.66) |
| Others † | 32 (5.03) |
| **Ethnic groups** |  |
| Wolof | 153 (24.02) |
| Fula | 139 (21.82) |
| Serer | 79 (12.40) |
| Jola | 16 (2.50) |
| Malinke | 17 (2.67) |
| Soninke | 9 (1.41) |
| Mauri | 7 (1.1) |
| Others † | 18 (2.82) |
| **Accommodation type** |  |
| Halls of residence | 138 (21.67) |
| Family home | 479 (75.19) |
| **Family members** |  |
| **Family members** |  |
| 1-2 | 37 (5.81) |
| 3-5 | 247 (38.77) |
| 6-8 | 188 (29.51) |
| 9 or plus | 74 (11.61) |
| **Matrimonial status** |  |
| Single | 558 (87.60) |
| Married | 60 (9.42) |
| Divorced | 2 (0.31) |
| **Smoker** |  |
| Yes | 10 (1.88) |
| No | 538 (84.30) |
| Stopped > 1 year | 10 (1.41) |
| **Alcohol consumer** |  |
| Yes | 21 (3.62) |
| No | 554 (86.81) |
| Stopped > 1 year | 3 (0.31) |
| **BMI Categorized** |  |
| Underweight | 69 (10.83) |
| Normal weight | 240 (37.68) |
| Overweight /Obesity | 53 (8.32) |
| **Blood Group** |  |
| A- | 9 (1.41) |
| A+ | 140 (21.98) |
| AB- | 1 (0.15) |
| AB+ | 25 (3.92) |
| B- | 5 (0.78) |
| B+ | 102 (16.01) |
| O- | 24 (3.77) |
| O+ | 300 (47.08) |
| **COVID-19 Vaccination** |  |
| Yes | 221 (35.01) |
| No | 365 (57.46) |
| Prefer not to say | 51 (7.54) |

N number; BMI Body Mass Indice; TASO technicians, administrative and service officers
† For nationality and ethnic group, others correspond to others of African nationality or ethnic group with very small numbers (maximum 5)

### 3.2. Compliance with preventive measures and antibodies’ seroprevalence

Most respondents reported a low level of compliance with the following preventive measures recommended by the health authorities: 31.39% among participants declared that they had worn a mask always or often, and 18.21% observed a physical distance (minimum 2 m) over the last 15 days. However, most participants reported a high level of compliance with hand washing: 75.35% had washed their hands always or often (Figure 2A). For social distancing, the majority of participants, 71.58%, reported 0-2 frequencies of Participation in social events during the 15 days before the survey; 70.8% declared that they used public transport at least 2 times a day; and finally, 62.16% had visited someone 15 days before the survey (Figure 2B).

**Figure 2:**
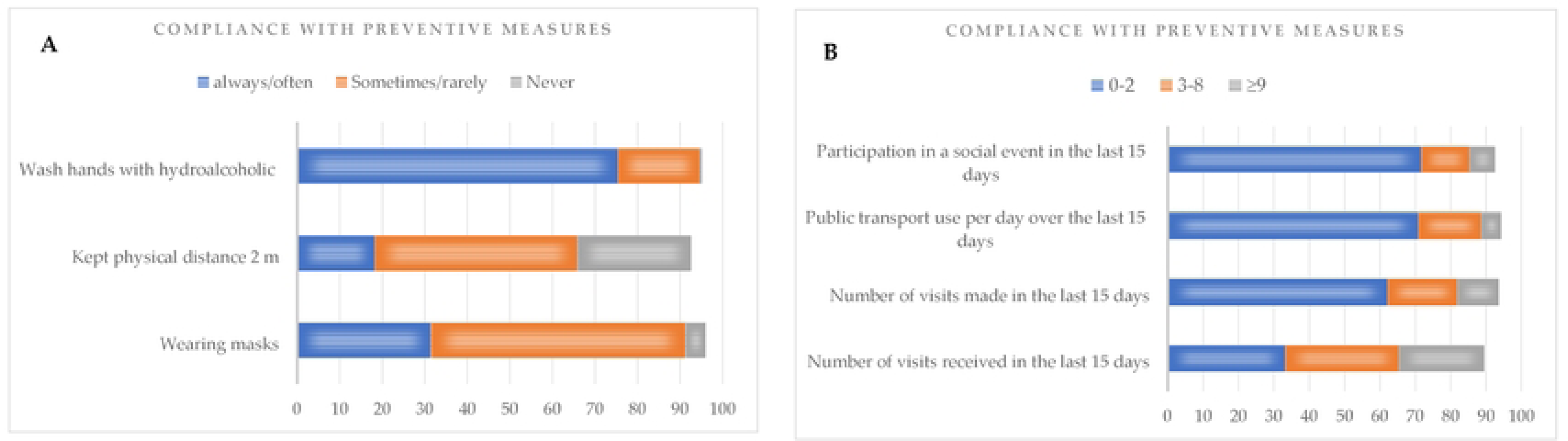
Compliance,,vith Preventive measures recommended by authorities

### 3.3. Seroprevalence of IgM et IgG SARS-CoV-2 antibodies

For overall participants, 6.91% (95% CI: 4.93 – 8.87) were seropositive for SARS-CoV-2 IgM antibody, and 92% (95% CI: 89.90 – 94.11) were seropositive for SARS-CoV-2 IgG. Then, we analysed IgM and IgG seroprevalence according to the different characteristics of the study population shown in Table II.

**Table II:**
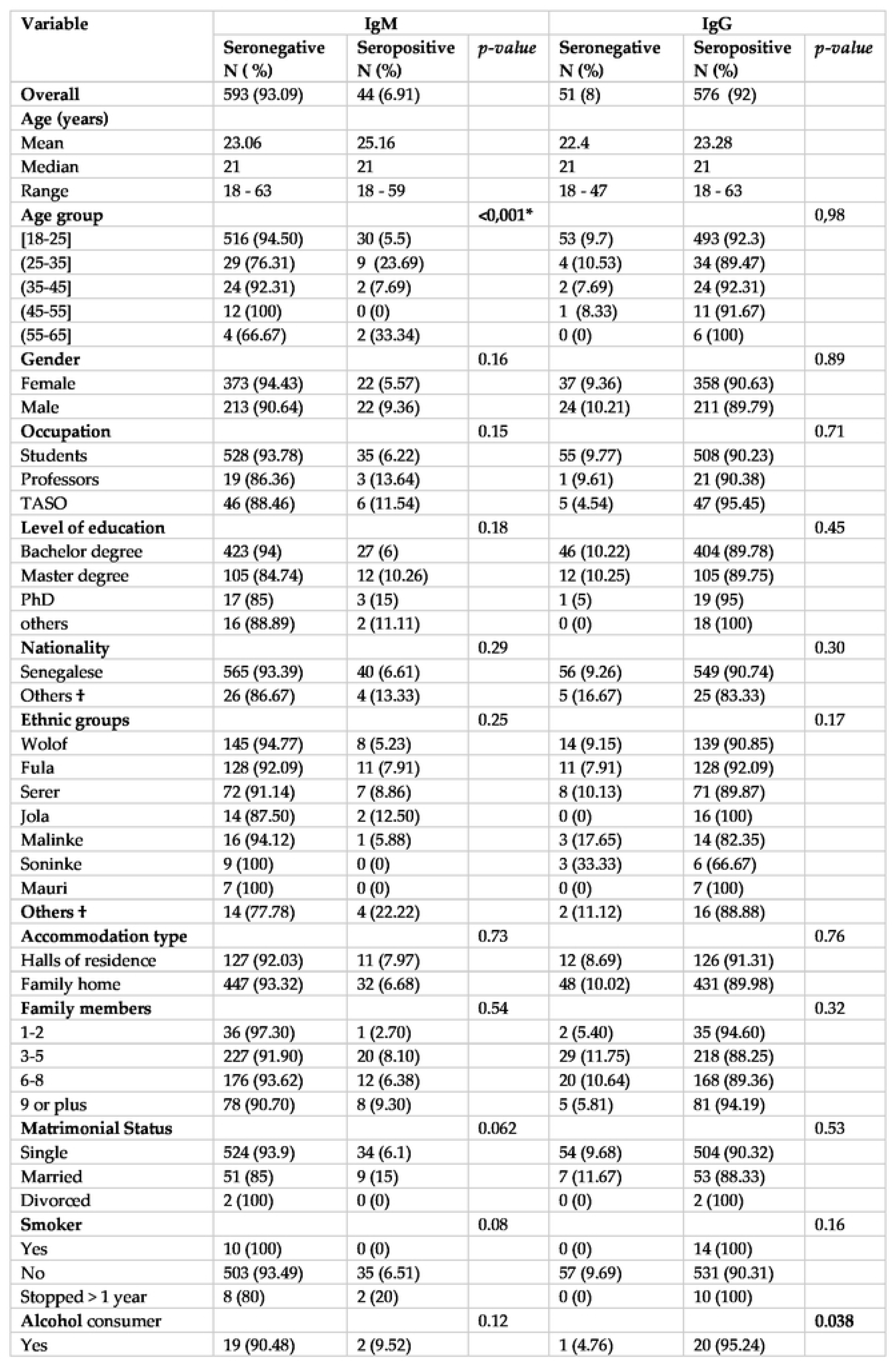
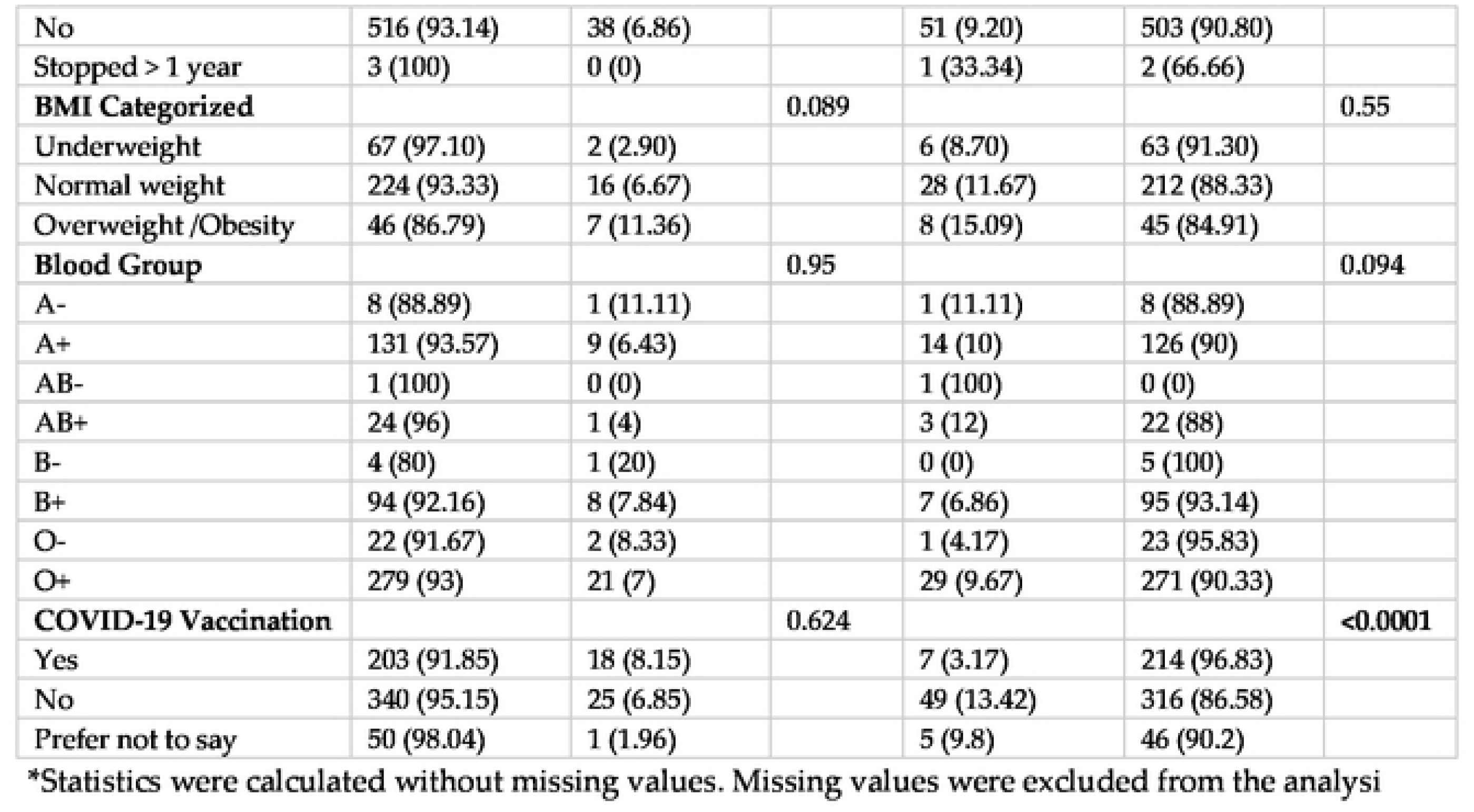
Distribution of SARS-CoV-2 lg Mand G seropositive and seronegative individuals among the 637 participants enrolled.

| Variable | IgM |  |  | IgG |  |  |
| --- | --- | --- | --- | --- | --- | --- |
|  | Seronegative<br>N ( %) | Seropositive<br>N (%) | *p-value* | Seronegative<br>N (%) | Seropositive<br>N (%) | *p-value* |
| **Overall** | 593 (93.09) | 44 (6.91) |  | 51 (8) | 576 (92) |  |
| **Age (years)** |  |  |  |  |  |  |
| Mean | 23.06 | 25.16 |  | 22.4 | 23.28 |  |
| Median | 21 | 21 |  | 21 | 21 |  |
| Range | 18 - 63 | 18 - 59 |  | 18 - 47 | 18 - 63 |  |
| **Age group** |  |  | <0,001* |  |  | 0,98 |
| [18-25] | 516 (94.50) | 30 (5.5) |  | 53 (9.7) | 493 (92.3) |  |
| (25-35] | 29 (76.31) | 9 (23.69) |  | 4 (10.53) | 34 (89.47) |  |
| (35-45] | 24 (92.31) | 2 (7.69) |  | 2 (7.69) | 24 (92.31) |  |
| (45-55] | 12 (100) | 0 (0) |  | 1 (8.33) | 11 (91.67) |  |
| (55-65] | 4 (66.67) | 2 (33.34) |  | 0 (0) | 6 (100) |  |
| **Gender** |  |  | 0.16 |  |  | 0.89 |
| Female | 373 (94.43) | 22 (5.57) |  | 37 (9.36) | 358 (90.63) |  |
| Male | 213 (90.64) | 22 (9.36) |  | 24 (10.21) | 211 (89.79) |  |
| **Occupation** |  |  | 0.15 |  |  | 0.71 |
| Students | 528 (93.78) | 35 (6.22) |  | 55 (9.77) | 508 (90.23) |  |
| Professors | 19 (86.36) | 3 (13.64) |  | 1 (9.61) | 21 (90.38) |  |
| TASO | 46 (88.46) | 6 (11.54) |  | 5 (4.54) | 47 (95.45) |  |
| **Level of education** |  |  | 0.18 |  |  | 0.45 |
| Bachelor degree | 423 (94) | 27 (6) |  | 46 (10.22) | 404 (89.78) |  |
| Master degree | 105 (84.74) | 12 (10.26) |  | 12 (10.25) | 105 (89.75) |  |
| PhD | 17 (85) | 3 (15) |  | 1 (5) | 19 (95) |  |
| others | 16 (88.89) | 2 (11.11) |  | 0 (0) | 18 (100) |  |
| **Nationality** |  |  | 0.29 |  |  | 0.30 |
| Senegalese | 565 (93.39) | 40 (6.61) |  | 56 (9.26) | 549 (90.74) |  |
| Others † | 26 (86.67) | 4 (13.33) |  | 5 (16.67) | 25 (83.33) |  |
| **Ethnic groups** |  |  | 0.25 |  |  | 0.17 |
| Wolof | 145 (94.77) | 8 (5.23) |  | 14 (9.15) | 139 (90.85) |  |
| Fula | 128 (92.09) | 11 (7.91) |  | 11 (7.91) | 128 (92.09) |  |
| Serer | 72 (91.14) | 7 (8.86) |  | 8 (10.13) | 71 (89.87) |  |
| Jola | 14 (87.50) | 2 (12.50) |  | 0 (0) | 16 (100) |  |
| Malinke | 16 (94.12) | 1 (5.88) |  | 3 (17.65) | 14 (82.35) |  |
| Soninke | 9 (100) | 0 (0) |  | 3 (33.33) | 6 (66.67) |  |
| Mauri | 7 (100) | 0 (0) |  | 0 (0) | 7 (100) |  |
| **Others †** | 14 (77.78) | 4 (22.22) |  | 2 (11.12) | 16 (88.88) |  |
| **Accommodation type** |  |  | 0.73 |  |  | 0.76 |
| Halls of residence | 127 (92.03) | 11 (7.97) |  | 12 (8.69) | 126 (91.31) |  |
| Family home | 447 (93.32) | 32 (6.68) |  | 48 (10.02) | 431 (89.98) |  |
| **Family members** |  |  | 0.54 |  |  | 0.32 |
| 1-2 | 36 (97.30) | 1 (2.70) |  | 2 (5.40) | 35 (94.60) |  |
| 3-5 | 227 (91.90) | 20 (8.10) |  | 29 (11.75) | 218 (88.25) |  |
| 6-8 | 176 (93.62) | 12 (6.38) |  | 20 (10.64) | 168 (89.36) |  |
| 9 or plus | 78 (90.70) | 8 (9.30) |  | 5 (5.81) | 81 (94.19) |  |
| **Matrimonial Status** |  |  | 0.062 |  |  | 0.53 |
| Single | 524 (93.9) | 34 (6.1) |  | 54 (9.68) | 504 (90.32) |  |
| Married | 51 (85) | 9 (15) |  | 7 (11.67) | 53 (88.33) |  |
| Divorced | 2 (100) | 0 (0) |  | 0 (0) | 2 (100) |  |
| **Smoker** |  |  | 0.08 |  |  | 0.16 |
| Yes | 10 (100) | 0 (0) |  | 0 (0) | 14 (100) |  |
| No | 503 (93.49) | 35 (6.51) |  | 57 (9.69) | 531 (90.31) |  |
| Stopped > 1 year | 8 (80) | 2 (20) |  | 0 (0) | 10 (100) |  |
| **Alcohol consumer** |  |  | 0.12 |  |  | 0.038 |
| Yes | 19 (90.48) | 2 (9.52) |  | 1 (4.76) | 20 (95.24) |  |
| No | 516 (93.14) | 38 (6.86) |  | 51 (9.20) | 503 (90.80) |  |
| Stopped > 1 year | 3 (100) | 0 (0) |  | 1 (33.34) | 2 (66.66) |  |
| **BMI Categorized** |  |  | 0.089 |  |  | 0.55 |
| Underweight | 67 (97.10) | 2 (2.90) |  | 6 (8.70) | 63 (91.30) |  |
| Normal weight | 224 (93.33) | 16 (6.67) |  | 28 (11.67) | 212 (88.33) |  |
| Overweight /Obesity | 46 (86.79) | 7 (11.36) |  | 8 (15.09) | 45 (84.91) |  |
| **Blood Group** |  |  | 0.95 |  |  | 0.094 |
| A- | 8 (88.89) | 1 (11.11) |  | 1 (11.11) | 8 (88.89) |  |
| A+ | 131 (93.57) | 9 (6.43) |  | 14 (10) | 126 (90) |  |
| AB- | 1 (100) | 0 (0) |  | 1 (100) | 0 (0) |  |
| AB+ | 24 (96) | 1 (4) |  | 3 (12) | 22 (88) |  |
| B- | 4 (80) | 1 (20) |  | 0 (0) | 5 (100) |  |
| B+ | 94 (92.16) | 8 (7.84) |  | 7 (6.86) | 95 (93.14) |  |
| O- | 22 (91.67) | 2 (8.33) |  | 1 (4.17) | 23 (95.83) |  |
| O+ | 279 (93) | 21 (7) |  | 29 (9.67) | 271 (90.33) |  |
| **COVID-19 Vaccination** |  |  | 0.624 |  |  | <0.0001 |
| Yes | 203 (91.85) | 18 (8.15) |  | 7 (3.17) | 214 (96.83) |  |
| No | 340 (95.15) | 25 (6.85) |  | 49 (13.42) | 316 (86.58) |  |
| Prefer not to say | 50 (98.04) | 1 (1.96) |  | 5 (9.8) | 46 (90.2) |  |

\*Statistics were calculated without missing values. Missing values were excluded from the analysis

Regarding IgM seroprevalence, the results showed significant age-dependence (*p* < 0.001) with higher seroprevalence in the (25-35) age group with 23.69% (95% CI: 20.39 – 27) compared to (18-25) age group with 5.5% (95% CI: 3.73 – 7.27). According to gender, IgM seroprevalence was higher in men, 9.4% (95% CI: 7.13 – 11.67) than in women, 5.6% (95% CI: 3.51 – 7.38), but no significant differences were found by sex (*p* = 0.83). There were no significant differences according to the other sociodemographic parameters regarding IgM antibodies seropositivity, such as occupation, level of education, ethnic group, accommodation type and size, but with some differences between sub-groups. For example, depending on the type of residence, we found a higher IgM seroprevalence among those living in halls of residence, 7.97% (95% CI: 5.87 – 10.07) compared to those living in family homes, 6.68% (95% CI: 4.74 – 8.62)], *p* = 0.73.

Concerning IgG seroprevalence, we found a significant difference between people who drank alcohol [95.24% (95% CI: 93.58 – 96.89)] and those who didn’t [90.80%, (95% CI: 88.55, 93.04)], with *p* = 0.038. Then, a very significant difference was found according to vaccination status, with a high seropositivity rate in vaccinated people [96.83% (95% CI: 95.47 – 98.19)] compared to unvaccinated [86.58% (95% CI: 83.93 – 89.22)], *p* < 0.0001. No significant difference was found between IgG seroprevalence according to age groups, while IgG seroprevalence was lower in men [89.9% (95% CI: 87.60 – 92.24)] than in women [90.6% (95% CI: 88.33 – 92.87)], *p* = 0.98. Then, no significant differences were found by sex (*p* = 0.83). IgM and IgG seroprevalences were higher in Professors and TAS officers than in students, yet this difference was not statistically significant (*p* = 0.71).

Concerning antibodies’ seroprevalence according preventive measures, we found a significant difference in IgM seroprevalence according to the ‘number of visits made to someone’ during the 15 days before the survey (*p* = 0.008) (Table III). However, we found no significant difference in IgG seroprevalence and compliance with barrier measures.

**Table III.** Distribution of Sars-Cov-2 lg M and G seropositive and seronegative according to preventive measures.

|  | IgM |  |  | IgG |  |  |
| --- | --- | --- | --- | --- | --- | --- |
|  | Seronegative<br>n ( %) | Seropositive<br>n ( %) | *p-value* | Seronegative<br>n ( %) | Seropositive<br>n ( %) | *p-value* |
| **Wearing masks** |  |  | 0.17 |  |  | 0.73 |
| Always/Often | 183 (91.04) | 18 (8.96) |  | 19 (9.45) | 182 (90.55) |  |
| Sometimes/Rarely | 355 (93.42) | 25 (6.58) |  | 38 (10) | 342 (90) |  |
| Never | 30 (100) | 0 (0) |  | 4 (13.33) | 26 (86.67) |  |
| **Kept a physical distance of 2 m** |  |  | 0.35 |  |  | 0.33 |
| Always/Often | 110 (94.83) | 6 (5.17) |  | 8 (6.90) | 108 (93.10) |  |
| Sometimes/Rarely | 277 (91.42) | 26 (8.58) |  | 31 (10.23) | 272 (89.77) |  |
| Never | 161 (94.15) | 10 (5.85) |  | 21 (12.28) | 150 (87.72) |  |
| **Wash hands with hydroalcoholic or with soap and water** |  |  | 0.105 |  |  | 0.599 |
| Always/Often | 451 (93.96) | 29 (6.04) |  | 51 (10.62) | 429 (89.38) |  |
| Sometimes/Rarely | 108 (88.52) | 14 (11.48) |  | 10 (8.20) | 112 (91.80) |  |
| Never | 4 (100) | 0 (0) |  | 0 (0) | 4 (100) |  |
| **Number of visits received in the last 15 days** |  |  | 0.583 |  |  | 0.971 |
| 0-2 | 205 (93.61) | 14 (6.39) |  | 23 (10.50) | 196 (89.50) |  |
| 3-8 | 188 (92.16) | 16 (7.84) |  | 21 (10.29) | 183 (89.71) |  |
| ≥9 | 146 (94.80) | 8 (5.20) |  | 15 (9.74) | 139 (90.26) |  |
| **Number of visits made in the last 15 days** |  |  | 0.008* |  |  | 0.946 |
| 0-2 | 373 (94.20) | 23 (5.81) |  | 41 (10.35) | 355 (89.65) |  |
| 3-8 | 109 (87.20) | 16 (12.80) |  | 12 (9.60) | 113 (90.40) |  |
| ≥9 | 73 (97.33) | 2 (2.67) |  | 7 (9.33) | 68 (90.67) |  |
| **Public transport use per day over the last 15 days** |  |  | 0.878 |  |  | 0.668 |
| 0-2 | 419 (92.9) | 32 (7.1) |  | 44 (9.75) | 407 (90.25) |  |
| 3-5 | 105 (92.92) | 8 (7.1) |  | 14 (12.39) | 99 (87.61) |  |
| ≥9 | 33 (91.67) | 3 (8.33) |  | 3 (8.33) | 33 (91.67) |  |
| **Participation in a social event in the last 15 days** |  |  | 0.589 |  |  | 0.289 |
| 0-2 | 423 (92.76) | 33 (7.24) |  | 51 (11.18) | 405 (88.82) |  |
| 3-8 | 79 (90.80) | 8 (9.20) |  | 5 (5.75) | 82 (94.25) |  |
| ≥9 | 44 (95.65) | 2 (4.35) |  | 4 (8.70) | 42 (91.30) |  |

Significant values are in bold.

### 3.5. Risk factors associated with SARS-CoV-2 IgM and IgG seropositivity among PHS community

Multiple Logistic regression analysis didn’t show an association between IgG seropositivity and the factors studied. Interestingly, an association between IgM seropositivity and participants’ ethnic group [O.R. 0.723 95% CI: 0.49 – 0.96, (*p* = 0.046)] was reported (Table IV). Using a univariate analysis, we found a significant association between IgM seropositivity and marital status [O.R. 2.399 95% CI: 1.08 – 4.87 (*p* = 0.021)], and Body Mass Indice (BMI) categorised [O.R. 2.015 95% CI: 1.12 – 3.56, (*p* = 0.016)]. In addition, on one hand, we found a significant association between vaccination and IgG seropositivity [O.R. 4.741 95% CI: 2.25 – 11.65, (*p* < 0.001)]; on the other hand, no associations were observed between SARS-CoV-2 seropositivity and sex and age groups. Finally, no significant association were found between IgM and IgG seropositivity and preventive measures (Table V).

**Table IV:** Multiple logistic regression of sociodemographical risk factors affecting seropositivity.

| Variable | IgM |  |  |  | IgG |  |  |  |
| --- | --- | --- | --- | --- | --- | --- | --- | --- |
|  | Univariate OR (95% CI) | *p-value* | Multivariate OR (95% CI) | *p-value* | Univariate OR (95% CI) | *p-value* | Multivariate OR (95% CI) | *p-value* |
| Age | 1.034 [0.99 - 1.06] | 0.055 | 0.971 [0,74 -1,21] | 0,808 | 1.023 [0.98 – 1.081] | 0.345 | 0.974 [0.89 – 1.071] | 0.573 |
| Gender | 1.75 [0.943 - 3.25] | 0.073 | 0.776 [0,10 – 4.6] | 0.791 | 0.906 [0.53 - 1.578] | 0.728 | 0.913 [0.52 - 1.62] | 0.757 |
| Occupation | 1.652 [0.92 – 2.72] | 0.065 | 1.345[0,06 – 22.03] | 0.834 | 1.26 [0.69 – 2.75] | 0.508 | 2.545 [0.60 – 15.33] | 0.250 |
| Level of education | 1.526 [0.91 – 2.47] | 0.095 | 1.709 [0.31 – 9.47] | 0.532 | 0.973 [0.61 – 1.61] | 0.911 | 0.917 [0.48 – 1.73] | 0.790 |
| Nationality |  |  | 0.074 [0.003 – 2.21] | 0.083 | 1.96 [0.64 – 4.93] | 0.186 | 2.286 [0.49 – 7.82] | 0.225 |
| Ethnic groups | 0.985 [0.86 – 1.13] | 0.826 | 0.723 [0.49 – 0.96] | **0.046 \*** | 0.95 [0.84 - 1.07] | 0.426 | 0.967 [0.85 – 1.095] | 0.6 |
| Place of residence | 0.995 [0.72 – 1.23] | 0.740 | 0.562 [0.17 – 1.24] | 0.23 | 0.998 [0.80 – 1.27] | 0.992 | 0.981 [0.74 – 1.32] | 0.894 |
| Accommodation type | 1.21 [0.57 – 2.40] | 0.60 | 4.209 [0.91 – 21.62) | 0.067 | 1.17 [0.62 – 2.37] | 0.643 | 0.924 [0.41 – 2.23] | 0.854 |
| Family members | 1.133 [0.77 – 1.66] | 0.522 | 1.849 [0.78 – 4.78] | 0.175 | 1.138 [0.81 – 1.6] | 0.452 | 1.234 [0.83 – 1.85] | 0.296 |
| Maritus status | 2.399 [1.08 – 4.87] | **0.021 \*** | 4.44 [0.28 – 54.32] | 0.235 | 1.02 [0.47 – 2.68] | 0.956 | 1.135 [0.43 – 3.95] | 0.818 |
| Smoking | 1.004 [0.21 – 2.48] | 0.995 | 2.897 [0.087 – 36.8] | 0.457 |  |  |  |  |
| Alcohol | 1.064 [0.18 - 3.17] | 0.926 | 2.06 [0.066 – 30.84] | 0.622 | 0.835 [0.32 – 3.25] | 0.747 |  |  |
| BMI Categorized | 2.015 [1.12 – 3.56] | **0.016 \*** | 1.215 [0.39 – 3.73] | 0.729 | 0.785 [0.49 – 1.29] | 0.329 | 0.718 [0.43 – 1.21] | 0.206 |
| Blood Group | 1.011 [0.89 – 1.152] | 0.861 | 1.049 [0.786 – 1.47] | 0.235 | 1.023 [0.91 – 1.14] | 0.679 | 0.988 [0.86 – 1.24] | 0.842 |
| Vaccination Status | 1.206 [0.63 – 2.25] | 0.560 | 0.428 [0.08 – 1.89] | 0.285 | 4.741 [2.25 – 11.65] | **<0.001 \*** | 2.714 [0.59 – 48.48] | 0.325 |

OR odds ratio. Signifcant values are in bold.

**Table V:** Multiple logistic regression of preventive measures affecting seropositivity.

| Variable | *IgM* |  | *IgG* |  |
| --- | --- | --- | --- | --- |
|  | Univariate<br>OR (95% CI) | *p- value* | Univariate<br>OR (95% CI) | *p- value* |
| Wearing masks | 0.796 [0,5893 - 1,073] | 0.134 | 0.966 [0.74 – 1.25] | 0.793 |
| Failure to respect the 2m distance | 0.980 [0,7380 - 1,278] | 0.886 | 1.229 [0.96 – 1.59] | 0.108 |
| Number of visits received in the last 14 days | 0.901 [0.701 – 1.15] | 0.404 | 1.002 [0.82 – 1.23] | 0.984 |
| Number of visits made in the last 14 days |  |  |  |  |
| Wash hands with hydroalcoholic | 0.854 [0.62 – 1.17] | 0.327 | 0.891 [0.68 – 1.17] | 0.405 |
| Public transport use per day over the last 15 days | 1.043 [0.75 – 1.40] | 0.793 | 1.072 [0.82 – 1.441] | 0.630 |
| Participation in a social event in the last 15 days | 0.952 [0.71 – 1.24] | 0.734 | 1.094 [0.86 – 1.420] | 0.473 |

## 4. Discussion

This is the first sero-epidemiological survey of COVID-19 carried out in an educational establishment in Senegal. The study aimed to determine the seroprevalence of IgM and IgG SARS-CoV-2 antibodies and associated risk factors in order to assess the extent of virus exposure within the Polytechnic High School in Cheikh Anta Diop University. The survey was carried out on samples collected from personnel and students from May 2022 to August 2022, after the fourth wave of COVID-19 in Senegal. A total of 637 participants were included in the study, 62% of whom were women and 37% men. The median age was 21, which is justified by the fact that the study was conducted in a university environment. Consequently, the study population predominantly comprises students representing around 86% of the participants.

We showed high IgG and IgM SARS-CoV-2 antibody seroprevalences of 92% (95% CI: 89.90 – 94.11) and 6.91% (95% CI: 4.93 – 8.87), respectively. The IgM seroprevalence indicates that the virus was circulating within PHS at the time of the survey, with people who were currently or recently infected. Indeed, in seroprevalence surveys, it is recommended to use IgM and IgG detection to diagnose recent and previous infections [32]. IgM antibody levels rise around a week after the initial infection, indicating a recent infection [22]. IgG antibodies appear later than IgM antibodies (generally within 14 days of infection). They can persist for up to a year [33, 34], meaning that IgG antibodies serve as an indicator of an old infection [35].

The very IgG high seroprevalence could be due to increased exposure to the SARS-CoV-2 virus and the anti-COVID vaccination policy implemented by the authorities. Data from the Ministry of Health showed that our study occurred after the fourth wave of COVID-19 in Senegal in January 2022, and the deployment of the COVID-19 vaccine began in February 2021 [36], which could explain our result. The first seroprevalence surveys carried out in Senegal were conducted in 2020 after the first wave and showed a lower seroprevalence of around 20%. Between June and September 2020, Seck *et al.* found a SARS-CoV-2 seroprevalence rate of 21.1% (95% CI = 16.7–26.1%) [37]. During the same period, Ahouidi *et al*. conducted a study between June and October 2020. They found that the overall positivity rate was 20.4% and significant geographical differences in seropositivity with a higher seroprevalence in Dakar, around 40% [38]. Then, Talla *et al.* found an adjusted national prevalence of anti-SARS-CoV-2 IgG/IgM in Senegal estimated to be 28.4% between October 24 and November 26, 2020 [39]. A high SARS-CoV-2 seroprevalences and its variability between localities were also reported in the second national population based cross-sectional survey (72,2%) conducted in November 2021 [40] and among pregnant women attending the antenatal consultation (69,4%) during the second and third waves of COVID-19 in Senegal [41].

Results showed a sharp increase in seroprevalence since 2020 in Senegal, and data corroborating the finding that several variants have circulated in the country, including the delta variant, which has been the most deadly and caused a peak in the number of confirmed cases in July 2021 [42]. Study carried out in Africa at the same time as our survey showed similarly high seroprevalence, with 74% between July and August 2022 in Bangui, Central African Republic [43]. In Nigeria, in early 2021, there was a high SARS-CoV-2 seroprevalence of 72.4% [44]. Given this situation, it seems very important to carry out cyclical epidemiological survey studies to measure the extent of the spread of the SARS-CoV-2.

Interestingly, we found a higher IgM seroprevalence in men than women (9.4% vs. 5.6%). Then, our result would suggest that men are more sensitive to infection and/or reinfection with SARS-CoV-2 than women. A study conducted at the start of the pandemic by Takahashi *et al*. showed that men were more susceptible to severe forms of COVID-19 than women [45]. Gender differences in immune responses were also reported, with higher levels of pro-inflammatory cytokines (IL-8 and IL-18) and a more robust induction of non-classical monocytes in men [46]. This is consistent with studies that analysed the sex difference in antibody responses [47]. In further studies, it will be interesting compare immune responses according to gender and serological status in order to discover the mechanisms underlying sex differences.

Our results also showed a different distribution by age group, with lower IgM seroprevalence in younger individuals, particularly those aged 18-25 years, compared to those aged 55-65 years (5.5% and 33.34%, respectively) with a *p* < 0,001. The 18-25 age group appears less affected by recent infections. This reinforces the data showing that young people are less affected by SARS-CoV-2 infection [48]. Furthermore, they often experience an asymptomatic or mild SARS-CoV-2 infection associated with lower serum titers [49]. In Senegal, during the first wave, it was reported that seroprevalence of SARS-CoV-2 was higher in patients older than 65 years [37].

We investigated whether there was a link between the type of residence and IgM seroprevalence to determine the proportion of new infections accordingly. Surprisingly, we didn’t find a significant difference in terms of IgM seroprevalence among those living in halls of residence [7.97% (95% CI: 5.87 – 10.07)] compared to those living in family homes [6.68% (95% CI: 4.74 – 8.62)], *p* =0.73. This invalidates our hypothesis that the particular ecosystem of CAD University, with its over 80,000 students from diverse origins, would constitute a niche for spreading the virus, which is not proven in our study. It also appears that, even though the PHS community is mainly be composed by young people, they do not seem to facilitate new infection, contrary to what has been reported in other studies [48, 50].

Concerning compliance with preventive measures, we found a low level of compliance with preventive measures recommended by the health authorities, particularly for wearing masks (31.40%) and keeping a physical distance (minimum 2 m) (17.42%). However, most participants reported high-level compliance with hand washing (75.35%). Social distancing and staying at home were also registered although to a lower degree. Our results were in contradiction with those of Kearney *et al.* who observed high compliance with recommended COVID-19 prevention behaviors among their sample of Senegal respondents, particularly for masking and personal hygiene practices [51]. This difference could be explained by the fact that our study population is younger and less inclined to comply with preventive measures [52, 53]. Our findings revealed that IgM seropositivity depends on the ‘number of visits made to someone’, meaning non-respect of social distancing (*p* = 0.008), which shows the importance of WHO measures concerning the adoption of extensive social distancing as a non-pharmaceutical intervention to reduce the infection’s spread and the associated mortalities [54]. However, studies have shown that the impact of social distancing measures depends upon a host of demographic, environmental, and economic dimensions [55].

Regarding vaccination, around 35% of participants had been partially or totally vaccinated, which is higher than the national vaccination rate of 16.9% at the moment of the survey [56]. This could be explained by our population’s education level and knowledge of vaccination’s stakes. Efforts were also made by the CAD University authorities, who set up a vaccination campaign at various sites during October 2021. However, the vaccination rate remains low compared to the national level, where more than 70% of the population vaccinated since the pandemic started [57].

Furthermore, we found no significant differences in IgM seroprevalences between vaccinated and unvaccinated individuals. It would, therefore, appear that vaccination does not protect against the new variants and reinfections, as has already been demonstrated in numerous studies [58–60] indeed, WHO recommendations suggest that seasonal vaccination is necessary to protect against new variants [61].

We investigated the association of sociodemographic, clinical and lifestyle factors, such as BMI, blood type, smoking and alcohol consumption, with seropositivity using logistics. Then, we found an association between BMI categorised and IgM seropositivity (O.R. 0.238, *p* = 0.043). Previous studies have shown that obese individuals (BMI > 30 kg/m²) were significantly more likely to be seropositive. It is uncertain whether the raised seroprevalence in these groups represents a greater risk of SARS-CoV-2 infection. However, obese individuals are known to experience more severe COVID-19 symptoms [62, 63].

Interestingly, a significant association between IgM seropositivity and ethnic group was found (O.R. 0.723, *p* = 0.046), suggesting a difference in susceptibility to SARS-CoV2-depending on race and ethnicity. According to previous findings, it seems there are disparities in the effects of COVID-19 infection among different racial/ethnic groups [64, 65]. Then, understanding the mechanisms for the disparity will help to evaluate the risk for COVID-19 according to ethnicity.

Our findings showed that IgM seropositivity was associated with marital status (unmarried/married) (O.R. 2.39, *p* = 0.021); another study reveals that transmission of SARS-CoV-2 is variable among different people within the home. For instance, the risk for infection was higher between spouses, at 43%, which could be a reflection of transmission through intimacy or longer or more direct exposure [66].

Regarding vaccination status, we found a relationship between vaccination and IgG seroprevalence (O.R. 4.741, *p* < 0.001), showing that people who had received vaccine doses were four times more likely to produce anti-SARS-CoV-2 antibodies than those who had not. It will be interesting to determine the IgG-neutralizing antibodies and to determine antibody levels according to vaccination status and type. Indeed, it has been shown that vaccines from different manufacturers might induce different antibody responses [67].

Some limitations of our study must be considered. By design, we carried out an on-site university population-based cross-sectional study. Thus, the results cannot be extrapolated directly to the general CAD University population. Participants were subjected to recall bias when completing the questionnaires, particularly preventive measures at the time of contact. We also noted a very low level of participation from professors and TAS officers. This constitutes a bias in the statistical calculations. Missing data for certain criteria and non-prefer responders were a limitation for statistics. To date, our study is the only SARS-CoV-2 sero-epidemiological survey conducted in a university community in Senegal. Data on the Serological testing performed by measuring IgM and IgG immunoglobulins against the SARS-CoV-2 targeting peak S1 protein separately is a strength of our study. This allowed us to differentiate recent from previous infection. Finally, with our questionnaire we were able to collect a lot of data, which was useful in calculating associations with risk factors.

In summary, our analysis of more than 630 subjects from Polytechnic High School community members estimated the extent of exposure to SARS-CoV-2. Our study revealed a high seroprevalence from May–August 2022 following the fourth wave of COVID-19 in Senegal. Our results show that the majority of students and staff have already been exposed to SARS-CoV-2 and confirm the circulation of the virus (SARS-CoV-2) at the time of the survey as shown by the high IgM and IgG seroprevalence. The data show a link between seropositivity and various factors such as age, non-compliance with prevention measures. These results underline the importance of sero-epidemiological surveys to estimate the real impact of the COVID-19 pandemic and the disparities between populations in order to establish a profile of the transmission dynamics of the virus. In addition, these results may be essential for the CAD university in the event of the emergence of future waves in order to make appropriate decisions and put in place means of monitoring the evolution of the pandemic after the relaxation of social distancing measures and the implementation of a vaccination schedule at CAD University, which will serve as a basis for other universities in Senegal.

## Author Contributions

“Conceptualization, F.T. and A.A.M.D, C.M.N, M.D.; methodology, F.T., A.A.M.D., C.S.C.A.N., S.S., S.D.T, S.C., A.S., D.D., and M.N.M; software, F.T., C.S.C.A.N., I.D., D.D., K.K., I.D.; A.L.D, M.D..; validation, F.T., A.A.M.D., C.M.N., S.M.S.; formal analysis, F.T., K.K, C.S.C.A.N., I.D., D.D.; A.L.D., M.D.; investigation, sample collection and biobank management, F.T., A.A.M.D., M.N.M, C.S.C.A.N, D.D., S.C., S.S., S.D.T., A.S.; resources, F.T., A.A.M.D., S.M.S, M.D.; data curation, F.T., S.S., C.S.C.A.N., D.D.; writing—original draft prepa-ration, F.T.; writing—review and editing, F.T., A.A.M.D., MNM, MD, CMN, KK, ID, SMS; visualization, F.T, A.A.M.D.; supervision, F.T.,A.A.M.D., S.M.S.,M.D.,C.M.N; project administration, F.T.; funding acquisition, F.T., A.A.M.D., and C.M.N. All authors have read and agreed to the published version of the manuscript

## Funding

This research was funded by Polytechnique High School competitive research impulse fund.

## Institutional Review Board Statement

The study was conducted in accordance with the Decla-ration of Helsinki, and approved approved by the Senegalese National Ethics Committee for Research in Health (Reference number N°000043/MSAS/CNERS/SP, 28 February 2022).

## Informed Consent Statement

Informed consent was obtained from all subjects involved in the study.

## Data Availability Statement

Not applicable.

## Acknowledgments

The authors would like to thank the top management of the “Ecole Supéri-eure Polytechnique de Dakar” for financial support and all students, Technicians, Administrative, Service officer and professors’ members who voluntarily participated to the study.

## Conflicts of Interest

The authors declare no conflict of interest. The funders had no role in the design of the study; in the collection, analyses, or interpretation of data; in the writing of the manuscript; or in the decision to publish the results”.

## Notes

### Competing Interest Statement

The authors have declared no competing interest.

### Funding Statement

The author(s) received no specific funding for this work.

